# Dynamic Profiling and Prediction of Antibody Response to Booster Inactivated Vaccines by Microsample-driven Biosensor and Machine Learning

**DOI:** 10.1101/2024.01.25.24301760

**Authors:** Sumin Bian, Min Shang, Ying Tao, Pengbo Wang, Yankun Xu, Yao Wang, Zhida Shen, Mahamad Sawan

## Abstract

Knowledge on the antibody response to inactivated vaccines in third dose is crucial because it is one of the primary global vaccination programs. This study integrated microsampling with optical biosensors to profile neutralizing antibodies (NAbs) in fifteen vaccinated healthy donors, followed by application of machine learning to predict antibody response at given timepoints. Over a nine-month duration, microsampling and venipuncture were conducted at seven individual timepoints. A refined iteration of fiber optic-biolayer interferometry (FO-BLI) biosensor was designed, enabling rapid multiplexed biosensing of NAbs towards both wild-type and Omicron variants in minutes. Findings revealed a strong correlation (Pearson r of 0.919, specificity of 100%) between wild-type NAbs levels in microsamples and sera. Following the third dose, Sera NAbs levels for wide-type increased by 2.9-fold after seven days and 3.3-fold within a month, subsequently waning and becoming undetectable in three months. Considerable but incomplete escape of the latest omicron subvariants from booster vaccine elicited NAbs was confirmed, although a higher number of binding antibodies (BAbs) was identified by another rapid FO-BLI biosensor in minutes. Significantly, FO-BLI highly correlated with a pseudovirus neutralization assay in identifying neutralizing capacities (Pearson r of 0.983). Additionally, machine learning demonstrated exceptional accuracy in predicting antibody levels with an error of <5% for both NAbs and BAbs across multiple timepoints. Microsample-driven biosensing enables individuals to access their results within hours after self-collection, while precise models could guide personalized vaccination strategies. The technology’s innate adaptability positions its potential for effective translation in diseases prevention and vaccines development.

## 1. Introduction

Antibody response to inactivated vaccines in a third dose (In-Vx-3) after two prime doses of inactivated vaccines has not been well investigated in the general population. This knowledge is crucial as the homologous inactivated vaccine booster is the primary vaccination program in China and many other countries. Given the possibility of recurring waves of COVID-19 outbreaks every six months, the potential for millions to be infected cannot be underestimated [1]. As we know, BA.4/5 and BF.7 are the dominant strains during the December wave of 2022 in China, and XBB.1.5 is a highly infectious subvariant that drove a new wave of infections in early 2023. How In-Vx-3 works towards the latest omicron variants is less well studied. Few studies showed that In-Vx-3 boosted the antibody response in individuals who had a low antibody response after the initial two doses and increased the level of neutralizing antibodies (NAbs) by up to five times in the general population [2, 3]. More research is needed to evaluate the immune persistence of the homologous In-Vx-3 and the antibody response towards the latest variants.

The NAbs levels highly correlate with immune protection from the SARS-CoV-2 infection [4] 4. However, the standard NAbs assays, including the pseudovirus neutralization test (PVNT) [5], have limitations because they require strict operating environment and well-trained personnel to perform complex steps. To address these issues, we designed a rapid and automated fiber optic-biolayer interferometry (FO-BLI) biosensor to detect NAbs in human sera with clinical suitability [6]. Nonetheless, it is uncertain whether the NAbs levels obtained through this biosensor precisely reflect the observed neutralization capacities in conventional assays. Moreover, venipuncture, a standard sampling method in hospitals, may not be ideal for frequent blood collection because it places additional burden and risk on healthcare systems and individuals. Meanwhile, microsampling has been established as a simple and painless method for antibody evaluation [7−10]. However, its utilization to evaluate NAbs is limited due to technical challenges, and subsequent measurements continue to use the traditional assays, which delays decision making. Therefore, clinical practice necessitates a rapid biosensor that matches the performance of a standard assay for multiplexed biosensing of NAbs towards both the wide-type (WT) strain and latest subvariants. A microsample-driven biosensor will be particularly valuable during a pandemic due to its non-invasive and high-throughput characteristics to understand vaccines efficacy. In addition to monitoring antibody levels, it’s crucial to predict these levels before sample measurements. This approach allows for individualized vaccination planning, which, in turn, facilitates proactive measures to prevent potential outbreaks. From this perspective, machine learning (ML) has been used to predict the antibody response following the second and third doses of mRNA vaccine in transplant recipients [10, 11]. However, both studies primarily focused on identifying the clinical predictors of the antibody response instead of predicting the antibody levels or their evolution over time in an individual.

To address these challenges, we integrated microsample dried blood spot (DBS) with the multiplexed FO-BLI biosensor to dynamically profile NAbs levels in 15 In-Vx-3 healthy donors. Subsequently, we used ML algorithms to predict the antibody levels for the same group of donors at various timepoints, using a dataset that included physiological parameters and optical measurements. Scheme 1 explains the study design in detail. Using the FO-BLI NAb biosensor, we further evaluated the extent to which the latest variants escaped neutralizing antibodies against wide type (WT-NAbs) by In-Vx-3. We also developed a simplified FO-BLI biosensor for the multiplexed detection of binding antibodies (BAbs) towards different lineages to gain a richer insight into the antibody response. Originally planned to last three months after booster dose, the study continued for six additional months, during which six donors provided samples on Day 270. This process enabled us to understand how the relaxed epidemic control measures, followed by the outbreak, affected the antibody response. This study paves the way for an individualized vaccination strategy and public health decision making. Also, the protocol of this integrated technology is well-established and highly adaptable, making it readily applicable for real-world uses in diseases prevention and vaccines evaluation across various contexts.

**Scheme 1.**
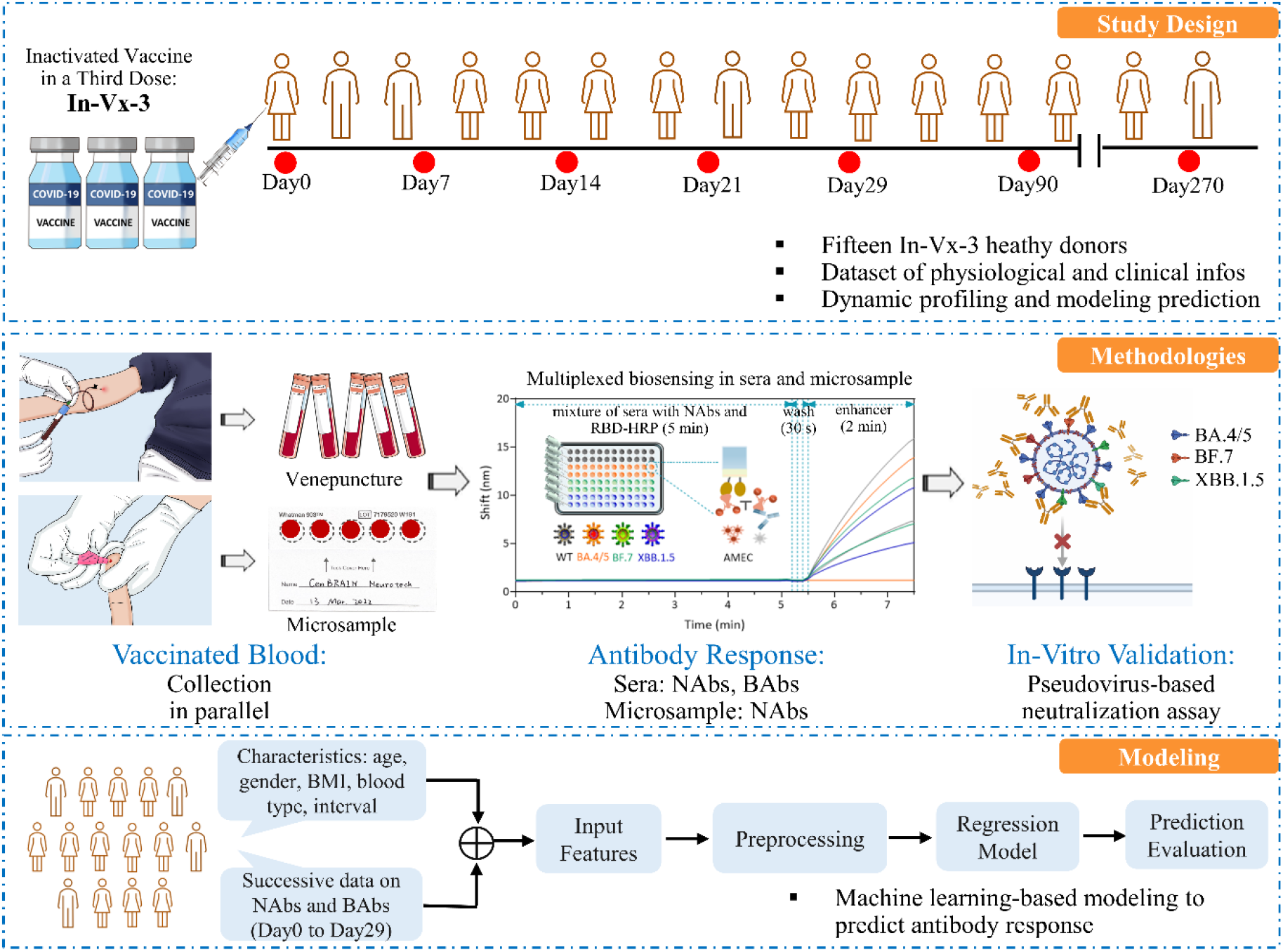
Scheme of the study design. The study involves recruiting fifteen healthy donors who received booster inactivated vaccine, collecting blood in parallel, conducting the multiplexed biosensing of the antibody response, validating the results using an in vitro pseudovirus-based neutralization test, and applying ML to predict the antibody response in individuals.

## 2. Materials and Methods

All the experimental sections are listed in detail in the ‘Supplementary Materials” file, including the following ten independent items: S-2.1. Materials and Equipment, S-2.2. Designing the FO-BLI biosensor for multiplexed detection of NAbs, S-2.3. Evaluating the fibers’ reproducibility for FO-BLI NAbs biosensor, S-2.4. Establishing DBS method for NAbs evaluation, S-2.5. Description of the study cohort, S-2.6. Designing the FO-BLI biosensor for multiplexed detection of BAbs in sera, S-2.7. Defining the limit of detection in both sera and DBS for NAbs, S-2.8. Validation of the FO-BLI NAbs biosensor using in vitro PVNT, S-2.9. Machine learning-based prediction of the antibody levels and S-2.10. Statistical Analysis.

## 3. Results

### 3.1 Multiplexed FO-BLI biosensor to detect NAbs in sera and microsamples

The multiplexed FO-BLI biosensor to detect NAbs towards WT and Omicron subvariants in 100-fold diluted sera was summarized in Table S1. Compared to its previous version [6], this biosensor used a more environmentally and user-friendly biomaterial 3-Amino-9-ethylcarbazole (namely, AMEC) as signal enhancer (adopted from our previous study [12]) and the high-salt SD buffer for sample dilution (adopted from another study [13])). Using functionalized fibers, multiple samples or one sample towards multiple subvariants were detected within 6.5-7.5 min (Fig. 1A). This protocol confirmed that bebtelovimab (BEB), a potent and fully human NAbs [14], was ineffective against XBB.1.5, despite its broad neutralizing activity towards omicron variant lineages BA.4/BA.5 and BF.7 (S-2.2; Fig. S1). Also, AMEC-precipitated fibers did not undergo complete regeneration (Fig. S2), resulting in a one-fiber-one-sample approach for measuring NAbs.

**Fig. 1.**
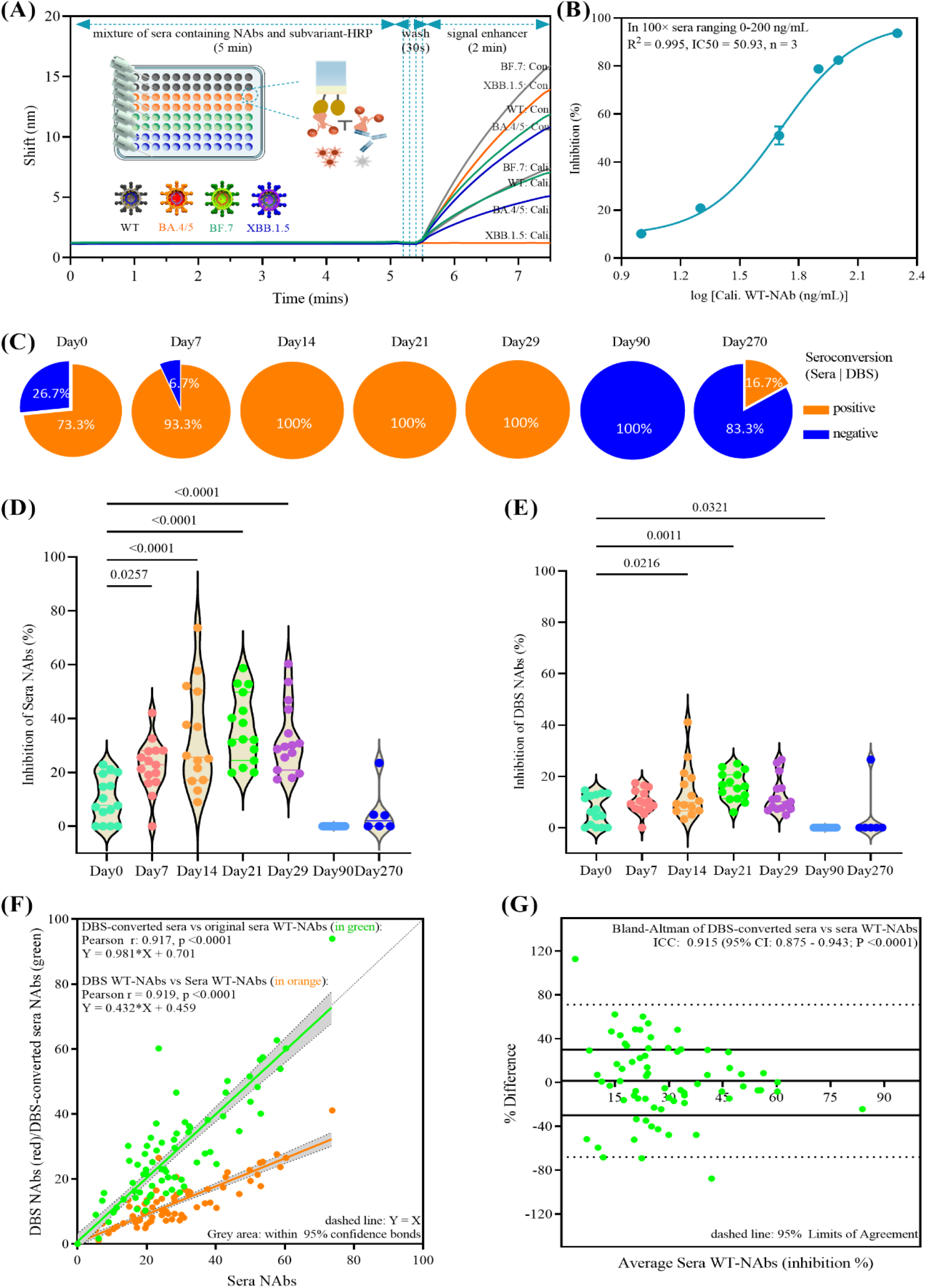
The multiplexed FO-BLI NAbs biosensor and long-term NAbs response towards wide type in the 15 enrolled In-Vx-3 donors in sera and microsamples. (A) Flowchart of the FO-BLI biosensor for multiplexed detection of a calibrator antibody as compared to a control. (B) Inhibition curve to detect the calibrator WT-NAbs in both matrices. (C) Profiling of WT-NAbs levels from Day0 till Day270; (D) WT-NAbs in sera; (E) WT-NAbs in paired microsamples; (F) Correlation between Sera vs DBS WT-NAbs (n = 94, data in orange) and DBS-converted vs original sera WT-NAbs (data in green); (G) Bland-Altman plot of relative difference between DBS-converted sera vs original sera WT-NAbs (n = 94).

When evaluating the DBS microsamples for NAbs biosensing following a former procedure [15], five out of the ten listed extraction buffers were discarded due to their high background interference (data not shown), and the other five remained for further consideration: Superblock T20, PBS+T80, PBS+0.5%BSA, PBS+0.05%T20 and PBS+0.05%T20+2%BSA. Further experiments proved that PBS+0.05%T20+2%BSA was the most suitable buffer, as the two tested artificial samples had similar performance compared to their sera counterpart (Table S2 (A) and (B)), showing the highest inhibition (mean of 75.2%, CV of 6.3%, n=4) and the lowest background (mean of 2.7%, CV of 8.9%, n=2). Using a series of calibrator WT-NAbs concentrations for spiking and extraction, the overall extraction efficiency was determined to be 80.9% with a CV of 5.4% (Table S2 (C)), indicating that 80.9% of calibrator WT-NAbs were extracted from DBS samples. Simultaneously, data confirmed the appropriateness to determine DBS concentrations by interpolating from the inhibition curve of the FO-BLI NAbs biosensor in 100-fold sera (Fig. 1B). Moreover, the LoDs for WT-NAbs detection in sera and DBS were determined to be 4.9% and 0.0%, respectively, using paired samples of six vaccine-naïve healthy donors.

### 3.2 Baseline characteristics of the study cohort

Table 1 shows the baseline characteristics of this cohort study, which consisted of 15 In-Vx-3 healthy donors, 73.3% of whom were female, the mean age was 25.0 y (means ± standard deviation (STD): 3.6), and the mean body mass index (BMI) was 19.9 (STD: 3.5). All participants received the third dose of COVID-19 vaccine in March 2022 with a mean interval of 7.1 months (STD: 1.10 between dose2 and dose3), and 86.7% received the inactivated Sinovac vaccine. Furthermore, 6 of 15 donors (40%) provided samples up to nine months after the third dose (i.e., Day 270), which coincided with the period of relaxed pandemic control measures and a local outbreak, where all donors resided. Consent of the enrolled donors and related approval from the ethics committees were received.

**Table 1.** Baseline characteristics of the study cohort. Overview characteristics of 15 In-Vx-3 healthy donors who received the third dose of COVID-19 vaccine with a follow-up duration of up to nine months

| No. | Sex | Age range | Blood type | BMI | Dose 1, 2, and 3 vaccines | Interval (month) | Last time point |
| --- | --- | --- | --- | --- | --- | --- | --- |
| 1 | Female | 21-25 | A | 19.9 | Sinovac | 8.3 | Day90 |
| 2 | Male | 21-25 | O | 28.7 | Sinopharm | 6.4 | Day90 |
| 3 | Female | 26-30 | AB | 19.8 | Sinovac | 6.9 | Day 270 |
| 4 | Female | 21-25 | B | 18.6 | Sinovac | 7.0 | Day90 |
| 5 | Female | 31-35 | O | 24.7 | Sinovac | 5.8 | Day90 |
| 6 | Female | 21-25 | A | 18.1 | Sinovac | 6.8 | Day270 |
| 7 | Female | 21-25 | O | 16.8 | Sinovac | 6.1 | Day270 |
| 8 | Male | 31-35 | B | 21.1 | Sinovac | 7.9 | Day90 |
| 9 | Female | 21-25 | O | 19.1 | Sinovac | 6.3 | Day90 |
| 10 | Female | 26-30 | O | 16.6 | Sinovac | 7.2 | Day270 |
| 11 | Female | 21-25 | O | 20.6 | Sinopharm | 6.5 | Day90 |
| 12 | Female | 26-30 | O | 19.9 | Sinovac | 9.6 | Day90 |
| 13 | Female | 26-30 | O | 20.9 | Sinovac | 8.0 | Day270 |
| 14 | Male | 26-30 | A | 20.1 | Sinovac | 9.0 | Day270 |
| 15 | Male | 21-25 | AB | 27.5 | Sinovac | 7.7 | Day90 |

### 3.3 Dynamic profiling of WT-NAbs in both sera and microsamples

Out of 94 clinical samples, the WT-NAbs levels in sera had 5.3-73.7% inhibition, with 25 samples falling the LoD. Samples that had WT-NAbs levels below a LoD of 4.9% were defined as negative with values set as zero. Particularly, the samples prior to the booster (i.e., Day0 sera) had a seropositivity of 73.3% (Fig. 1C) and a mean WT-NAbs of 7.7% (STD: 8.4%; Fig. 1D). We observed a 2.9-fold increase (STD: 9.7%) in the overall levels of sera WT-NAbs one week after vaccination. This increase was slightly higher in Day 14 samples at 3.3-fold (STD: 19.1%) and Day 21 samples at 3.6-fold (STD: 12.8%), which reached a seropositivity of 100%. The increase remained at 3.2-fold (STD: 13.1%) after one month but subsequently gradually declined, and WT-NAbs became undetectable after three months. Notably, the liberalization of epidemic control measures elected WT-NAbs in only one of the six remaining volunteers with a level of 23.5%. Compared to venous sera, DBS microsample sensitively monitored WT-NAbs with 100% specificity and identified an identical seropositivity for all samples of Days 0−270 (Fig. 1C). Of all 94 samples, DBS also revealed 25 samples that failed the LoD, while the remainder had DBS WT-NAbs levels of 1.2–41.1% of inhibition. Samples prior to the booster dose (i.e., Day 0 DBS) had a mean WT-NAbs of 6.2% (STD: 5.8%), and a 1.6-fold increase (STD: 10.4%) in the mean levels of sera WT-NAbs was found on Day 7 (Fig. 1E). This increase was relatively stable for Day 14 samples at 1.4-fold (STD: 10.4%) and Day 21 samples at 1.6-fold (STD: 5.6%). The increase remained at 1.3-fold (STD: 7.0%) after one month and gradually declined to become undetectable after three months. Similarly, WT-NAbs was detected in the paired DBS sample of the same volunteer who elicited serum antibody on Day 270.

Collectively, the Pearson correlation analysis revealed a strong correlation between DBS and Sera WT-NAbs (Pearson r = 0.919, p <0.0001, n = 94; Fig. 1F, in orange). A linear regression equation ([WT-NAbs]DBS = [WT-NAbs]sera *0.432 + 0.459, R^2^ = 0.919, p < 0.0001) was found to convert the DBS WT-NAbs levels to sera levels (i.e., DBS-converted sera). The WT-NAbs levels of the DBS-converted sera were not significantly different from the originally obtained sera levels (correlation coefficient (ICC) = 0.917, P < 0.0001) (Fig. 1F, in green). The Bland-Altman plot revealed an average bias of 1.5 (95% CI: 68 – 71%) between DBS-converted and originally-obtained sera WT-NAbs levels, where 74% of samples were within 30% of their mean (Fig. 1G). These results satisfy the criteria of FDA and EMA, which require at least 67% of clinical samples to be within 30% of their mean of difference.

### 3.4 Multiplexed profiling of NAbs towards Omicron BA.4/5, BF.7 and XBB.1.5

The study further assessed the extent to which the newest omicron lineages could evade neutralization by WT-elicited antibodies. At the start of the study WT coronavirus was predominant, however, as of early 2023, BA.4/BA.5, BF.7 and XBB.1.5 lineages have become more prevalent. The extended duration of the study has rendered DBS extractions collected at the beginning stages less suitable for antibody evaluation. Therefore, sera samples were used for this evaluation. Results showed that nine (9.6%) out of 94 sera exhibited positive NAbs activity towards BA.4/BA.5, five (5.3%) were positive towards BF.7, and one (1.0%) was positive towards XBB.1.5 (Table S3). These data suggest considerable but incomplete escape of BA.4/BA.5, BF.7, and XBB.1.5 from the WT-NAbs elicited by the third dose. Notably, of six Day 270 sera collected after the relaxed epidemic control measures that caused a significant Omicron outbreak, three sera simultaneously developed NAbs towards BA.4/BA.5 (50%) and BF.7 (50%), and one serum (of donor Nr. 13) reacted towards XBB.1.5. This result shows the broad-spectrum characteristics of this serum NAb towards Omicron subvariants.

### 3.5 Validation of the FO-BLI NAbs biosensor using clinically validated PVNT

It is critical to understand whether the NAbs levels obtained through this multiplexed NAbs FO-BLI biosensor precisely reflect the neutralization capacities observed in standard assays. To investigate this issue, a commercially available and clinically validated PVNT was performed to determine the IC50 of three sera towards the latest subvariants. Fig. 2(A−C) shows the inhibition percentage profiles of three sera NAbs and their IC50 values determined by the PVNT. These profiles demonstrate consistent positivity and negativity towards each subvariant. Comparison of IC50 values by the PVNT and NAbs levels by the FO-BLI NAbs biosensor reveals a strong correlation (Fig. 2D; Table S4) with a Pearson r of 0.983 (P <0.0001) and a linear regression of Y = 0.082*X-0.711. Of note, S1 is the serum collected from donor Nr. 11 on Day14, S2 is the serum collected from donor Nr. 10 on Day 10, and S3 is the serum from donor Nr. 13 on Day 270 as named in Table S3.

**Fig. 2.**
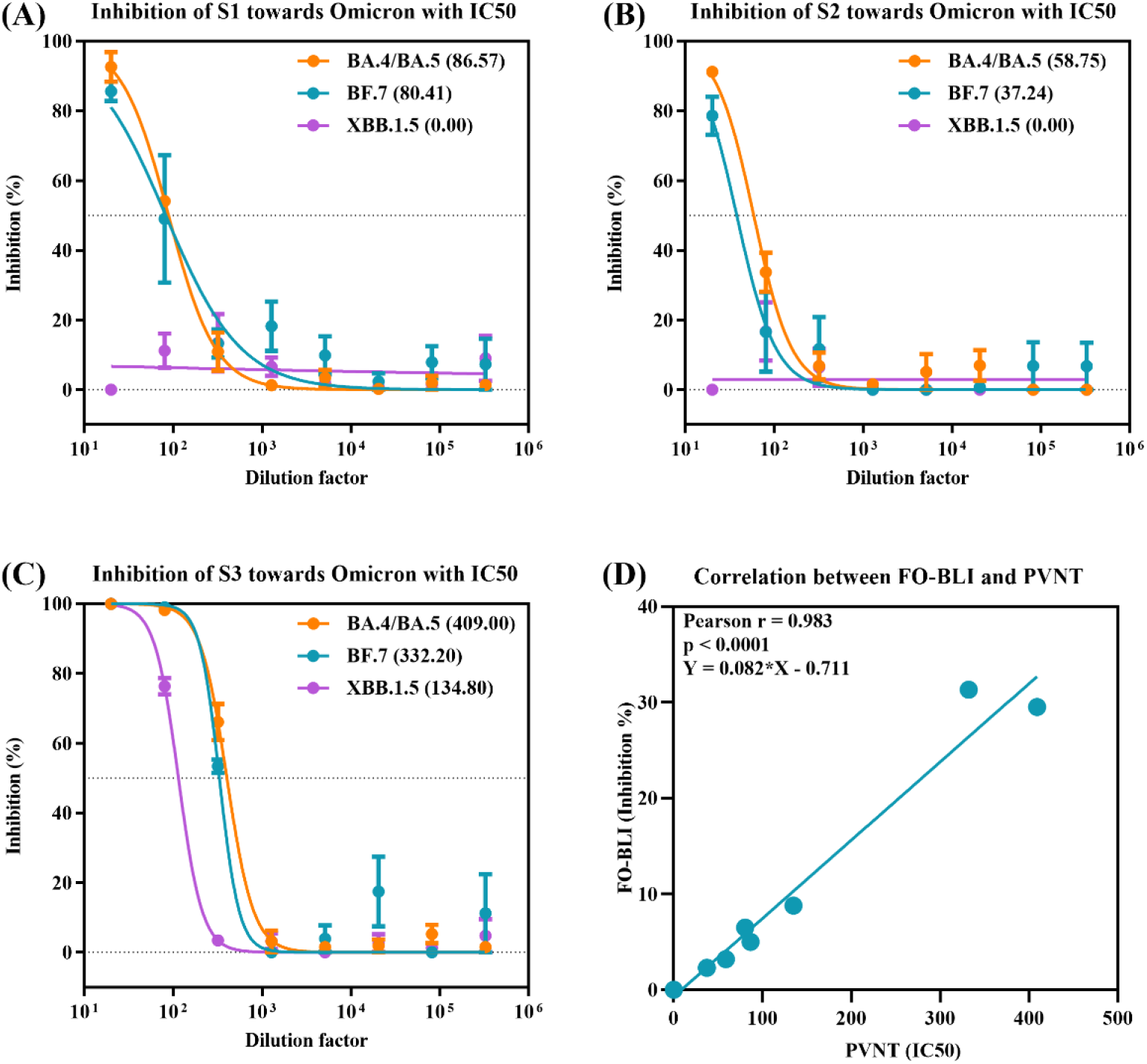
Validation of the FO-BLI NAbs biosensor using a PVNT. (A-C) Inhibition percentage profiles with IC50 values of three serum NAbs (S1-S3) towards omicron BA.4/BA.5, BF.7 and XBB.1.5, as measured using the clinically validated PVNT. (D) Correlation of the measurements between FO-BLI and PVNT.

### 3.6 Multiplexed biosensing of serum BAbs towards both WT and variants

A simplified version of the multiplexed BAbs FO-BLI biosensor was developed to obtain a more comprehensive profile of the antibody response of all donors (Fig. 3A). The signal-enhancer-free sensor is regenerable up to six times when using 10 mM Glycine pH 2.0 as the regeneration solution with a recovery of above 90.5% and a zero-baseline drift (Fig. 3B). Pre-functionalized probes allowed the detection of BAbs in 40-fold diluted sera in 7.0 mins (Table S5). Moreover, the FO-BLI BAbs biosensor demonstrated excellent reproducibility when tested on seven individual sera, with a less than 12% during two separate runs (Table S6). These results support the cost-effectiveness, robustness, and reliability of the BAbs biosensor for use in clinical samples. Besides, the FO-BLI BAbs biosensor provided additional evidence that the BEB antibody cannot bind XBB.1.5 despite its activity against previous subvariants (Fig. S3).

**Fig. 3.**
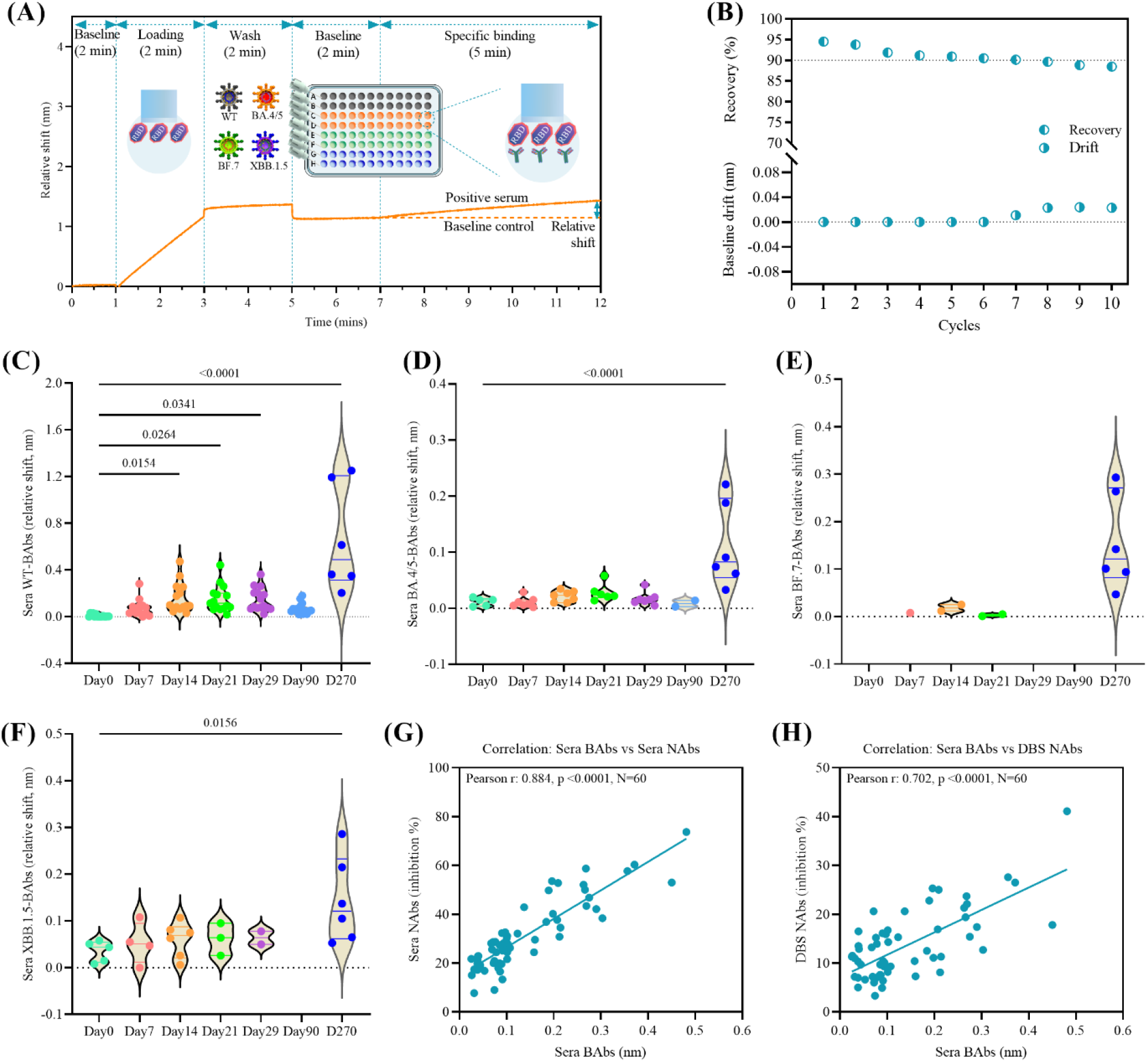
Performance of the multiplexed FO-BLI BAbs biosensor and long-term BAbs response in the 15 enrolled In-Vx-3 donors. (A) Flowchart of the multiplexed FO-BLI BAbs biosensor towards the WT strain and omicron subvariants in 40-fold diluted serum. (B) Regeneration capability of the FO-BLI BAbs biosensor using Glycine 10 mM pH 2.0 as the solution. Profiling of the sera BAbs levels towards (C) the WT strain, (D) BA.4/BA.5. (E) BF.7 and (F) XBB.1.5 on Days 0–270 using the FO-BLI BAbs biosensor. Correlation between (G) Sera BAbs versus Sera NAbs levels and (H) Sera BAbs versus DBS NAbs levels for Days 0–29 samples.

For the study cohort, BAbs levels by sera towards the WT strain, BA.4/BA.5, BF.7 and XBB.1.5 were profiled. The relative shifts of WT-BAbs in 94 sera exhibited significant variation, and nine samples fell below the LoD (see Fig. 3C). Moreover, we observed a notable 8.5-fold increase in overall levels of sera WT-BAbs one week after the vaccination. This increase was particularly striking on Day 14, where the levels increased by 20.7 folds, and remained stable on Day 21 (at 19.1-fold) and Day 29 (at 18.5-fold). However, three months after the vaccination, the increase in WT-BAbs dropped to 8.9-fold. Notably, following the liberalization of epidemic control measures, the levels of WT-BAbs sharply climbed to 82.9 folds on Day 270. High BAbs levels were developed towards the three omicron lineages with seropositivities of 42.5%, 11.7% and 27.6% for BA.4/BA.5 (Fig. 3D), BF.7 (Fig. 3E), and XBB.1.5 (Fig. 3F), respectively. Notably, all six Day 270 sera collected during the Omicron outbreak produced BAbs towards the three subvariants. Further analysis of the measurements in the first month revealed a clear correlation between the relative shifts of sera WT-BAbs and inhibition percentages of WT-NAbs in both sera (Pearson r = 0.884, Fig. 3G) and DBS (Pearson r = 0.702, Fig. 3H).

### 3.7 Machine learning-based modeling to predict the antibody levels over time

The section aims to investigate the feasibility of predicting the antibody response using ML techniques. Table S7 shows the initial performance of Sera NAbs, DBS NAbs and Sera BAbs obtained by six regression models to predict antibody levels at Day29. The Lasso Linear model outperformed all the other five models with a less than 5% RMSE and was therefore selected for further prediction takes. We then continued to investigate whether we could make a longer prediction based on relatively limited data. Thus, we predicted the measurements at a given time point (i.e., the *T*^*th*^ measurement) based on previous measurements (i.e., *p*M), where *T* ∈ {2, 3, 4}, *pM* ∈ {1, 2, 3}. Table 2 shows the prediction performance of a series of *T* and *pM* sets. According to Table 3, the Lasso linear model accurately predicted the measurements with an RMSE within 5% at several timepoints. Regarding the Sera NAbs, data from Day 0, 7 and 14 could predict the data on Day 29 with similar accuracy (RMSE of 3.7%). Similar accuracy was observed in the DBS NAbs. Interestingly, regarding DBS NAbs, Day 0 data could precisely predict the data of Day 7 (RMSE of 4.9%), and the data of Days 0−7 could well predict the Day 14 data (RMSE of 5.3%). Compared with the serum NAbs, DBS NAbs could be predicted with high precision at two more time points. Regarding the serum BAbs, more accurate data were predicted using this Lasso linear model. For example, data from Day 14 and onwards were successively obtained from the data of Days 0−7. Fig. 4(A−C) displays visual representations of the three parameters, where the predicted values are plotted against the actual measured values.

**Fig. 4.**
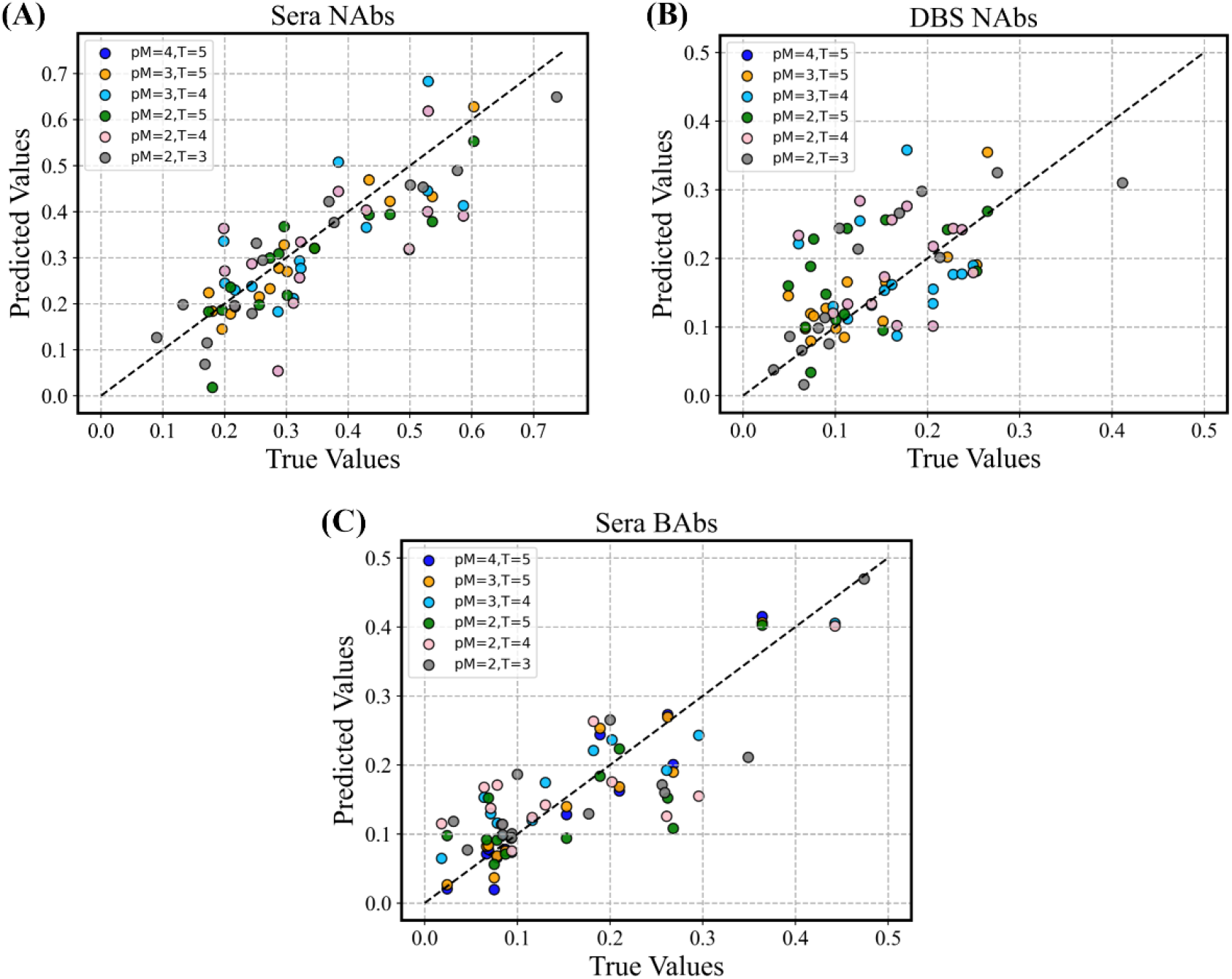
Visualization of the three different parameters. Profiles of three main parameters: predicted (A) serum NAbs, (B) DBS NAbs and (C) serum BAbs versus the true values.

**Table 2.** ML-modeling-based prediction. Prediction performance of the *T*^*th*^measurement (measurements at a given time point) based on previous measurements (*p*M). The prediction was made using the Lasso linear model for three parameters.

| Input of previous measurements ($pM$),<br>output at a given time point ($T$) | Sera NAbs<br>(RMSE) | DBS NAbs<br>(RMSE) | Sera BAbs<br>(RMSE) |
| --- | --- | --- | --- |
| $pM = 4, T = 5$ | 3.1% | 4.2% | 2.7% |
| $pM = 3, T = 5$ | 3.7% | 4.0% | 2.6% |
| $pM = 3, T = 4$ | 9.0% | 6.3% | 4.2% |
| $pM = 2, T = 5$ | 6.0% | 6.5% | 4.7% |
| $pM = 2, T = 4$ | 9.9% | 6.1% | 6.5% |
| $pM = 2, T = 3$ | 6.6% | 5.3% | 5.3% |
| $pM = 1, T = 5$ | 14.4% | 10.1% | 7.8% |
| $pM = 1, T = 4$ | 17.3% | 10.1% | 10.2% |
| $pM = 1, T = 3$ | 15.9% | 11.2% | 9.5% |
| $pM = 1, T = 2$ | 9.4% | 4.9% | 7.1% |

## 4. Discussion

This study integrated the multiplexed FO-BLI biosensor with microsampling for rapid and dynamic profiling of NAbs towards both WT strain and omicron variants in 15 In-Vx-3 healthy donors with a follow-up of nine months. We collected 94 paired samples and found that the levels of WT-NAbs from DBS microsamples were consistent with those obtained in venipuncture sera, with a Pearson correlation of 0.919 and a specificity of 100%. A linear regression was built to support a simple interchange of the data, proving the accuracy and reliability of DBS in evaluating NAbs under the optimized condition. Microsampling additionally proved its potential to meet the global demand for both large-scale and personalized management [16]. Moreover, validation of the FO-BLI NAbs biosensor using nine individual sera of three vaccinated donors showed its excellent accuracy in assessing neutralizing capabilities compared to a standard PVNT (Pearson r correlation 0.983). This further confirms the clinical utility of the biosensor and the reliability of the data produced to build ML models. To obtain a richer profile of antibody response, we simplified the FO-BLI BAbs biosensor to regenerate a single fiber for multiple uses and enable high-throughput screening at a lower cost.

With our technologies, the data showed a 3.3-fold increase in sera WT-NAbs levels after 0.5 months and a 3.2-fold increase after one month following the booster inactivated vaccines compared to the levels at Day 0. A similar trend was found with the DBS WT-NAbs levels (1.4-fold and 1.3-fold). However, the increase was sharper in WT-BAbs levels during the same period (20.7-fold and 18.5-fold). Despite the undetectable levels in WT-NAbs at three months, all sera developed good amounts of WT-BAbs (8.5-fold increase). After the epidemic control was loosened, the volunteers produced the elicited antibodies in both types, which were possibly restored by the Omicron outbreak. The antibody response towards Omicron, including its subvariants BA.4/BA.5, BF.7 and XBB.1.5, has shown extensive but incomplete escape from the WT-NAbs elicited by the booster. However, we identified a broad-spectrum serum NAb (Day 270 sample of donor 13) that could neutralize all of the latest Omicron subvariants, which offers a potent NAb candidate. Similarly, all six donors available on Day 270 during the Omicron outbreak developed more antibodies towards all three subvariants in both NAbs and BAb levels. Our data and others provide compelling evidence that a homologous booster of inactivated vaccine can enhance antibody response, although its advantages against omicron variants are limited [17]. Thus, there is a need in receiving a heterologous or an omicron-specific booster vaccine to better address the challenges posed by these variants [18, 19].

ML is poised to play a pivotal role in driving deep evidence-based medicine [20]. Although numerous clinical studies have employed ML to predict antibody response following vaccination, most have centered on predicting the probability of seroconversion at a single timepoint instead of predicting precise levels of antibody levels and their temporal dynamics in individuals [10, 11, 21−24]. Additional studies investigated the prediction of neutralization titers to multiple Omicron variants after breakthrough infection [25]. In this work, we bridged this gap by developing a model that can accurately predict the changes in antibody response at multiple timepoints based on previous measurements. Of note, our study found that simple linear models performed better than more complex ML algorithms, likely due to the limited size of our dataset. Moreover, we chose not to apply the model to predict antibody response toward Omicron, as the inactivated vaccines primarily target the ancestral strain, and the complex nature of emerging variants presents challenges for accurate modeling. This model aims to explore a valuable framework to predict the booster-induced antibody levels in healthy individuals, eventually enhancing the precision of vaccination strategies tailored to this specific population.

Our study does have several limitations. Firstly, the study cohort’s sample size is relatively small, which restricted our ability to fully comprehend the antibody response to the booster inactivated vaccine and construct a more precise prediction model. To enhance the model’s accuracy, further investigations are necessary, involving larger datasets that encompass a balanced representation across genders and age groups. Additionally, incorporating lifestyle-associated parameters extracted from microsamples would bolster these efforts [26]. Secondly, the microsamples collected during the early stages did not allow for the evaluation of NAbs against Omicron due to the extended duration of our study. This highlights the imperative need to enhance the storage stability of microsamples for prolonged research studies. Thirdly, despite the robust correlation observed between the FO-BLI NAbs biosensor and the pseudovirus-based neutralization assay, validating the findings using a real-virus neutralization assay holds paramount importance in a clinical context.

## 5. Conclusions

Collectively, we have developed two advanced FO-BLI biosensors that enable the rapid and multiplexed detection of NAbs and BAbs against the wild-type strain and omicron sublineages. The FO-BLI NAbs biosensor exhibits a strong correlation with a standard PVNT demonstrating its clinical utility for real-life application. The regenerable capability of the FO-BLI BAbs biosensor makes it a promising and cost-effective tool for high-throughput screening. Moreover, our use of microsampling as a simple yet reliable method for dynamically profiling NAbs has opened new avenues for personalized antibody response monitoring. Integrating microsample with the FO-BLI biosensor enables individuals to receive their test results within hours after self-sampling. The obtained accurate ML model in predicting antibody levels at multiple timepoints in the study cohort not only affirms the biosensor’s reliability but also underscores its potential to expedite decision-making for personalized vaccination strategies. Additionally, the system’s innate adaptability positions it favorably for addressing various disease prevention priorities and assessing novel vaccine candidates.

## Data Availability

All data produced in the present study are available upon reasonable request to the authors.

## CRediT authorship contribution statement

S. Bian: Conceptualization, Methodology, Experiments, Analysis, Writing, Reviewing, Editing, Funding acquisition. M. Shang: Recruiting volunteers, Collection of blood samples, Reviewing. Y. Tao: Methodology, Collection of samples, Experiments, Analysis. P.B. Wang: Collection of samples, Experiments; Y.K. Xu: ML Analysis, Writing; Y. Wang: Collection of samples; Z.D. Shen: Collection of samples; M. Sawan: Supervision, Reviewing, Editing, Funding acquisition.

## Data and materials availability

Data will be available on request.

## Acknowledgments

Authors appreciate all the volunteers for contributing their valuable samples and time for this study. This research was supported by the National Natural Science Foundation of China [Grant No. 82104122], the Research Center for Industries of the Future of Westlake University [Grant No. 210230006022219/001] and the Westlake University [Grant No. 10318A992001].

## Appendix A. Supplementary data

Supplementary data include Methods and Materials, Fig. S1-Fig. S3 and Table S1-S7.

## Supplementary Materials

### S-2. Methods and Materials

#### S-2.1. Materials and Equipment

SARS-CoV-2 related proteins were purchased from Sino Biologicals (Beijing, China), including Biotinylated ACE2 Protein Human Recombinant (His Tag, Cat 10108-H08H), SARS-CoV-2 (2019-nCoV) Spike RBD-His Recombinant Protein (Cat 40592-V08H), SARS-CoV-2 (BA.4/BA.5/BA.5.2) Spike RBD Protein (His Tag, HPLC-verified, Cat 40592-V08H130), SARS-CoV-2 BA.4.6/BF.7 (Omicron) Spike RBD Protein (His Tag, Cat 40592-V08H140), SARS-CoV-2 XBB (Omicron) Spike RBD Protein (His Tag, HPLC-verified, Cat 40592-V08H144). SARS-CoV-2 (2019-nCoV) Spike Neutralizing Antibody, Rabbit Mab (Cat 40592-R001) (namely, WT-R001) and Research Grade Bebtelovimab-DVV00319 (namely, BEB) were ordered from Sino Biologicals (Beijing, China) and AtaGenix (Wuhan, China), respectively. Biotinylation Kit/Biotin Conjugation Kit (Fast, Type B) - Lightning-Link^®^ (Cat ab201796), HRP Conjugation Kit - Lightning-Link^®^ (Cat ab102890) and Anti-Rheumatoid Factor IgM ELISA Kit (Cat ab178653) (Abcam, Shanghai, China); Tween 80, Tween 20 and Bovine Serum Albumin (BSA) (Sigma-Aldrich, Shanghai, China); SuperBlock™ (PBS) Blocking Buffer (Cat 37515) and SuperBlock™ T20 (PBS) Blocking Buffer (Cat 37516) (Thermo Scientific, Shanghai, China); ImmPACT* AMEC Red* Substrate Kit (Cat SK-4285; Vectorlabs, Shanghai, China); BD Microtainer^®^ contact-activated lancet (Cat 366594) (BD, Shanghai, China); WhatmanTM 903 Protein Saver Card (Cat 10534612) (Cytiva, Shanghai, China) and 96-well polystyrene black microplates (Greiner Bio-One GmbH, Shanghai, China). Streptavidin (SA, Cat 18-5020) sensors came from Sartorius Group (Gottingen, Germany). Sample diluent (SD) buffer [i.e., PBS (10 mM, pH 7.4) containing 0.02% Tween20 and 0.1% BSA] and high-salt SD buffer (i.e., SD containing 274 mM NaCl) were freshly prepared. Equipment includes an Octet^®^ K2 2-Channel System, a RED96E 8-Channel System (Sartorius Group, Gottingen, Germany), a Biotek ELx808 microplate reader (BioTek Instruments, USA) and a Axiocam 208 color (ZEISS, Oberkochen, Germany).

#### S-2.2. Designing the FO-BLI biosensor for multiplexed detection of NAbs

The multiplexed NAbs FO-BLI biosensor was adapted from a previous version (Bian et al., 2022) with two major improvements. First, the optical signal was enhanced using a more environment and user-friendly biomaterial 3-Amino-9-ethylcarbazole (namely, AMEC). AMEC-based biosensing resulted in a higher signal-to-noise ratio and the precipitates generated were soluble in alcohols, providing an opportunity for fiber regeneration (Tao, et al., 2022). Second, a high-salt SD buffer was used to dilute the serum 100-fold to ensure minimal interference from blood, as proved by another recent study of us (Bian et al., 2021). Similarly, the FO-BLI NAbs biosensor employed HRP-conjugated RBD to compete with NAbs present in the sample. If the HRP-RBD outcompeted NAbs in either affinity or amount, AMEC will bind with HRP to produce a good number of precipitates, thereby amplifying the signals in sub-seconds. In this study, NAbs were evaluated towards both WT and subvariants. Thus, WT-R001 continued to serve as the calibrator for WT-R001 while BEB was selected as the calibrator for Omicron NAbs. BEB is a potent and fully human NAbs with broad neutralizing activity to SARS-CoV-2 variants of concern, including Omicron variant lineages (Hentzien et al., 2022).

#### S-2.3. Evaluating the fibers’ reproducibility for FO-BLI NAbs biosensor

We assessed whether the precipitates induced by AMEC on the fiber tips could be entirely dissolved in ethanol, allowing for the fibers to be cleaned and regenerated. Evaluation was performed on 15 individual fibers, on which a mixture of a blank sample with HRP-WT was applied to interact with AMEC, resulting in final optical signals ranging from 0.5 to 40 nm, controlled by adjusting the reaction duration. The 15 precipitated fibers were divided into three groups. Group one was used to observe the surface morphology before any treatment, while group two and three were immersed in high-purity 99.75% ethanol at 4℃ for 2 hrs and overnight, respectively, to demonstrate the changes in surface morphology after treatment.

#### S-2.4. Establishing DBS method for NAbs evaluation

Fingerpick microsample-based dried blood spot (DBS) for NAbs determination was developed according to the protocol that we established for monoclonal antibody analysis (Bian et al., 2020). This study first evaluated the most appropriate buffer for NAbs detection using WT-R001 as the calibrator. First, WT-R001 (0 and 8 ug/mL) were spiked in a mixture of whole blood from healthy donors. Similarly, 40 uL of spiked blood was spotted on filter papers and air-dried for 1 h, followed by being punched out into a 6 mm diameter disc (approximately 10 uL of blood) and immersed into 240 uL of extraction buffer, resulting in a 25-fold predilution. In this study, ten different buffers were tested, including SuperBlock T20 buffer, SuperBlock buffer, SD buffer, high-salt SD buffer, PBS + 0.1% Tween80, PBS + 0.1% Tween20, PBS + 0.05% Tween20, PBS + 0.5% BSA, PBS + 0.05% Tween 20 + 2% BSA and PBS buffer. The following treatment with shaking, centrifugation and collection of supernatants were the same as in the former study. WT-R001 concentrations were measured using the FO-BLI NAbs biosensor. The buffer that contributed to the highest extraction efficiency of the two DBS samples was elected. Afterwards, WT-R001 with series concentrations of (0−1−2−5−10−20 ug/mL) were tested to carefully evaluate the precision of DBS for NAbs determination as compared to sera.

#### S-2.5. Description of the study cohort

This study builds upon our previous work on establishing the FO-BLI biosensor and was conducted with the ethic approval of both the Sir Run Run Shaw (SRRS) Hospital, School of Medicine Zhejiang University (research 20210706-7) and the Institutional Review Board of Westlake University (20210301BSM001). Informed consent was obtained from all the 15 In-Vx-3 healthy adult donors without a previously reported COVID-19 infection. The original dose could be Sinopharm or Sinovac, and the third dose could also be either of them. Paired samples were collected at seven defined time points: Day 0 (immediately prior to receiving dose 3), Day 7 (seven days after dose 3), Day 14, Day 21, Day 29, Day 90, and Day 270. Notably, Day 270 samples were provided by six participants who underwent relaxed epidemic control measures in China. Whole blood was collected via venipuncture into BD Vacutainer^®^ Lithium Heparin tubes and then centrifuged at 2000 rpm for 10 mins at 25°C to obtain the sera. Capillary blood was obtained from a finger prick and absorbed onto a protein saver card, with the first blood drop discarded. After air drying, a DBS was extracted. All samples were subsequently de-identified. Levels of NAbs in paired DBS and sera were detected using the FO-BLI NAbs biosensor. Moreover, physiological parameters of all participants were included in this study, such as age, gender, body mass index (BMI), blood type, and the interval between the third and second doses.

#### S-2.6. Designing the FO-BLI biosensor for multiplexed detection of BAbs in sera

A simplified version of our previously designed FO-BLI bioassay for multiplexed determination of binding antibodies (BAbs) in sera was developed (Bian et al., 2022). The purpose was to remove the use of signal enhancer to maximize the chance of fiber reuse without compromising the sensitivity. In summary, SA fibers were first immersed in SD buffer for 10 mins to establish a baseline measurement. Next, the fibers were functionalized with biotinylated WT-RBD to capture WT-specific BAbs in clinical samples. Biotinylated WT-RBD was diluted to 2 µg/mL in SD buffer, and the capture shifts were controlled to achieve approximately 1.2 nm. The detection of BAbs was carried out by directly dipping the functionalized fibers into clinical sera, which were first diluted by a factor of 40 to ensure an optimal signal-to-noise ratio, using the high-salt SD buffer. Healthy control sera from six vaccine-naïve volunteers were also used as negative controls to determine the LoD. The process for establishing the BA.4/BA.5-specific BAbs, BF.7-specific BAbs, and XBB.1.5-specific BAbs biosensors was identical.

#### S-2.7. Defining the limit of detection in both sera and DBS for NAbs

To define the limit of detection (LoD) of the techniques, we required a pool of healthy control serum that did not contain SARS-CoV-2 antibodies. Therefore, we recruited six healthy adult volunteers who had not received any vaccine for various reasons to donate paired blood. Volunteers were selected if their serum samples were tested negative for SARS-CoV-2 S-ECD, as determined using the Anti-S-ECD BAbs biosensor with signal enhancer (Bian et al., 2022), which was previously developed. Afterwards, paired sera and DBS were collected as healthy control serum and healthy control DBS to determine the LoD for sera and for DBS respectively. The same control sera were used to define the LoD for the new version of FO-BLI BAbs assay as well in the following section.

#### S-2.8. Validation of the FO-BLI NAbs biosensor using in vitro PVNT

The in vitro PVNT was conducted following a Nature protocol (Nie et al., 2020). Serum samples were serially diluted four-fold, starting from a 1:20 dilution, and then incubated with a SARS-CoV-2 pseudovirus luciferase reporter (GeneScript, SC2087A) at 37°C for 1 hr. The antibody-recombinant virus mixture was then added to Opti-HEK293/ACE2 cells. After 48 hrs., the cells were harvested, and infected cells were lysed. A luciferase chromogenic solution was added, and the luciferase luminescence (RLU) signals were measured using a microplate reader (Thermo, Variokan LUX) after incubation at room temperature for 3−5 mins. The percentage of infected cells was normalized to that derived from cells infected with SARS-CoV-2 pseudovirus luciferase reporter in the absence of serum. The neutralization percentage was calculated using equation (*Eq. 1*). ACE2-Fc was used as a positive control. Neutralization capacities were presented as 50% maximal inhibitory concentration (IC50). Same procedure was applied for the IC50 of three selected serum NAbs against BA.4/BA.5, BF.7, and XBB.1.5. Samples that did not reach 50% inhibition at the dilution of 1:20 were considered non-neutralizing and set at zero for correlation calculation.

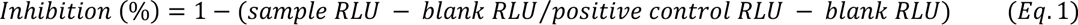

#### S-2.9. Machine learning-based prediction of the antibody levels

This section presents the implementation of ML-based inhibition prediction on three different parameters: Sera NAbs, DBS NAbs and Sera BAbs. The workflow is depicted in Fig. S1, including merging features, preprocessing, regression modeling, and prediction performance evaluation modules. Due to some absences on Day 270 and no inhibitions found after three months, we only considered the first five batches of measurements for ML-based prediction experiments. Additionally, two participants missed one measurement, leaving 13 participants for further study.

Our primary objective was to predict Day 29 inhibition using the first four datasets of Days 0−21. We considered the characteristics of each participant such as age, gender, blood type, BMI, and days to receive the third dose for ML. Prior to prediction modeling, we applied preprocessing techniques to each feature, including one-hot encoding for gender and blood type features, and normalization of age, BMI, and dose features to a range of 0 to 1. The first four inhibition results remained unchanged. Due to the similar inhibition trends of the three parameters, we combined the five characteristic features with the first four set of inhibition measurements from each test as input and used the 5th measurement (Day 29) from each test as the target output. Therefore, we extracted three samples from each participant, resulting in a total of 39 (13×3) samples. Finally, we constructed a regression-based prediction task with input features in a shape of 9 (4 measurements + 5 characteristics), and the target output as Day29 inhibition.

To identify the best regression model for prediction, we employed six different models, including two linear models, two polynomial models with varying degrees, support vector machine (SVM), and multilayer perceptron (MLP) methods (Jordan and Mitchell, 2015). We evaluated model performance using root mean square errors (RMSE) between predicted and true values. To validate our results, we implemented a leave-one-out validation scheme, where each run left one sample as the validation sample, and the remaining samples were used to train the regression model. This resulted in 39 RMSEs, which we divided into three groups according to three different parameters (i.e., 13 RMSEs for each parameter). We then computed the mean RMSE for each parameter. RMSE for each parameter was calculated as follows:

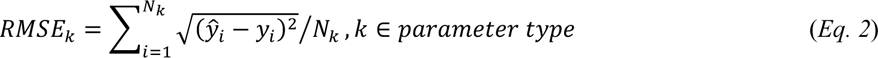

where, k is one of the 3 test parameters, and *N*_*k*_in this work should be 13 for each type of *k*. 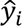 and *y*_*i*_ stands for the predicted target and the true target, respectively.

#### S-2.10. Statistical Analysis

P values for differences between groups of sera and DBS NAbs were determined using ANOVA (and nonparametric or mixed) methods for multiple comparisons and presented in violin plots, using GraphPad Prism v9.5.1 (GraphPad Software, California, USA). All the statistical tests were two-tailed, and P values less than 0.05 were considered statistically significant. Inhibition curves against RBD-NAbs and Omicron-NAbs were generated by the FO-BLI NAbs biosensor, and the half maximal inhibitory concentration (IC50) was determined using "dose-response-inhibitor: log(inhibitor) vs normalized response—variable slope". The in vitro PVNT principle in Fig. 1 was created by Biorender and ML visualization was drawn by Python 3.8.0. To assess the correlation of inhibition in paired sera and DBS, GraphPad Prism 9.5.1 was used, setting any value below the defined LoD as zero. Data are presented as means ± standard deviation (STD), as described in the corresponding figure legends. Linear regression lines were fitted to the data, and their 95% confidence intervals (CI) were calculated using GraphPad Prism. Relative Bland-Altman plots were created using GraphPad Prism, and the intra-class correlation coefficient (ICC) was determined using the "two-way mixed, absolute agreement—single measure test (absolute agreement)" from SPSS Statistics 25 (IBM, NY, USA) to investigate mean differences and agreement.

## S-3. Supplementary Results

### Supplementary Figures

**Fig. S1.**
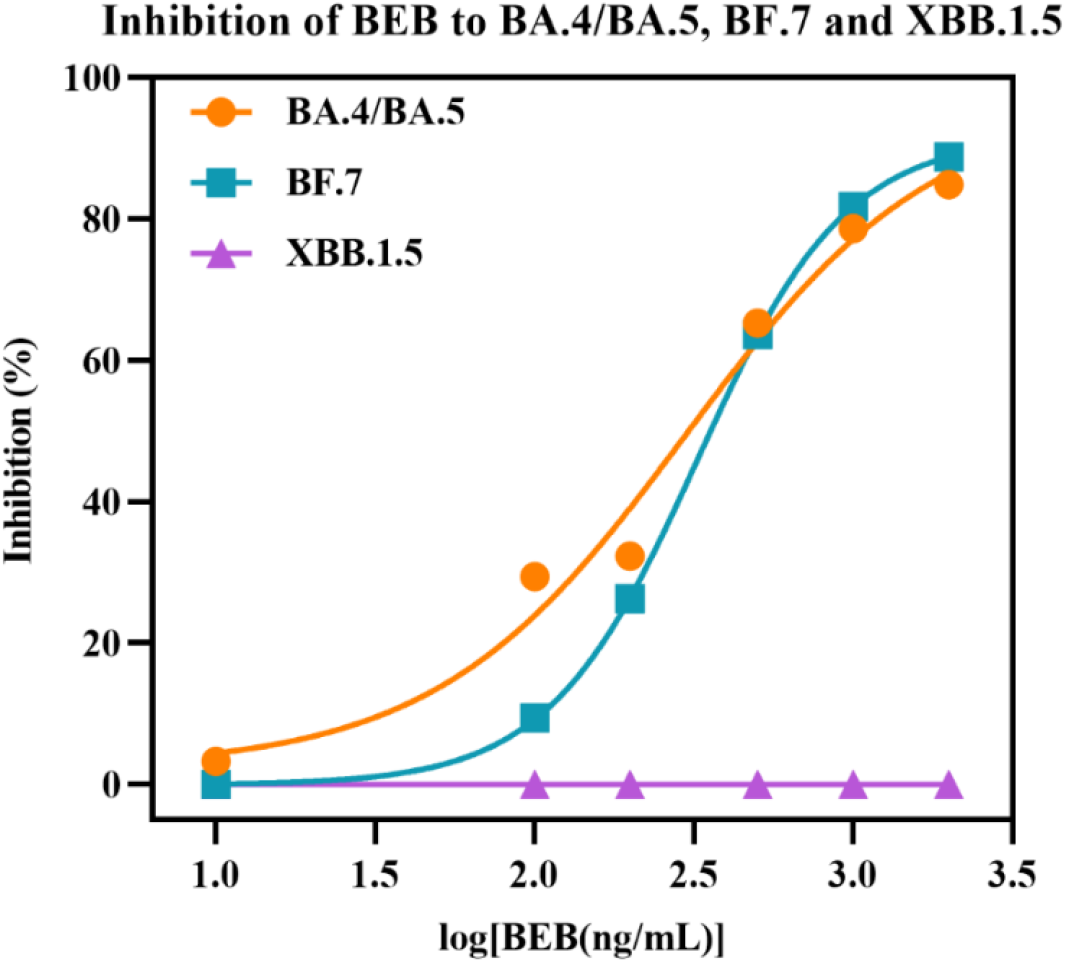
Inhibition of the calibrator BEB towards BA.4/BA.5, BF.7 and XBB.1.5 by the FO-BLI NAbs biosensor.

**Fig. S2.**
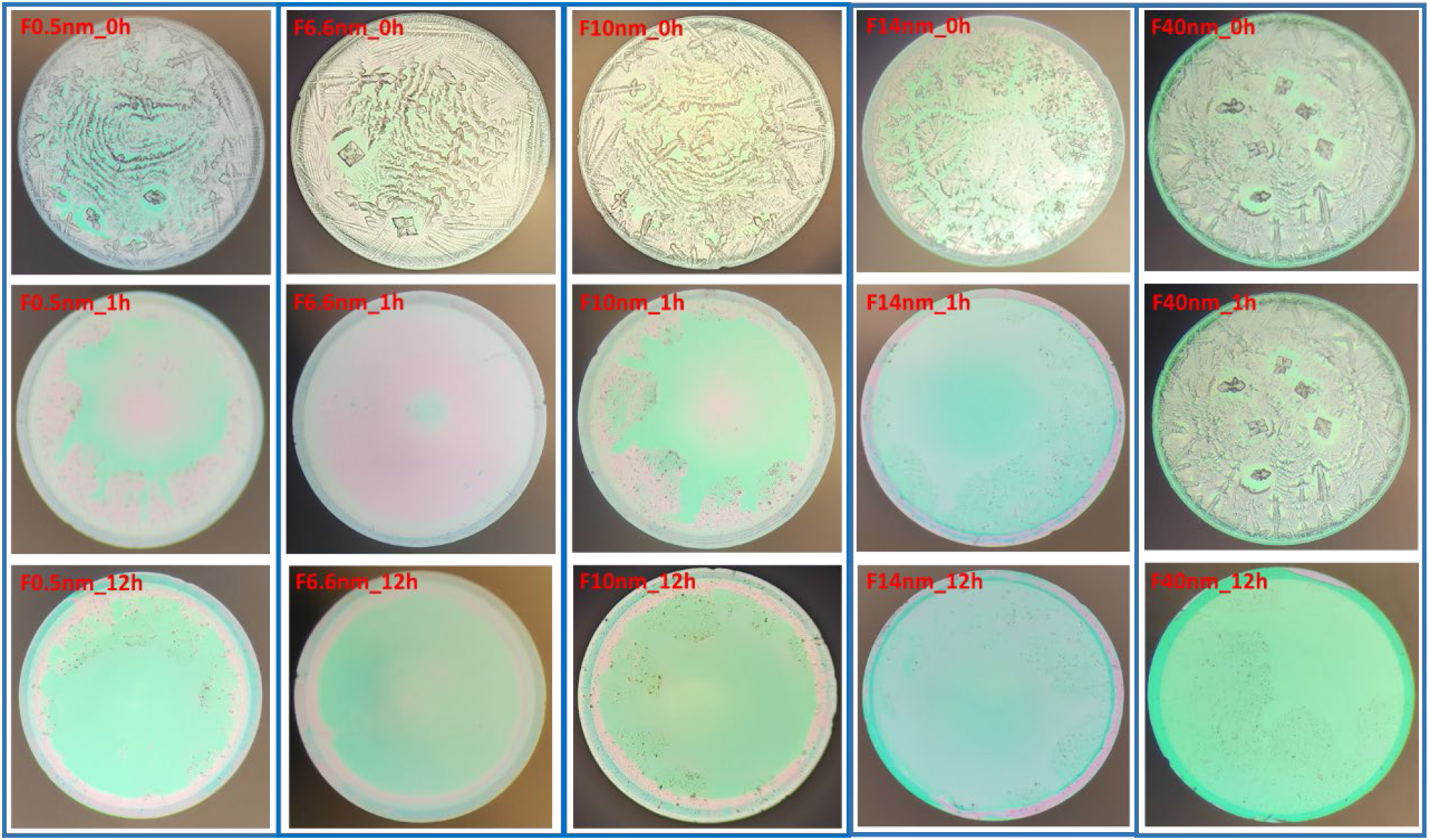
Three groups of fibers with controlled signals ranging from 0.5 nm to 40 nm were prepared to evaluate the effect of high-purity ethanol on cleaning and regenerating the fibers. Data showed that no fiber can be fully cleaned for reuse under the condition tested.

**Fig. S3.**
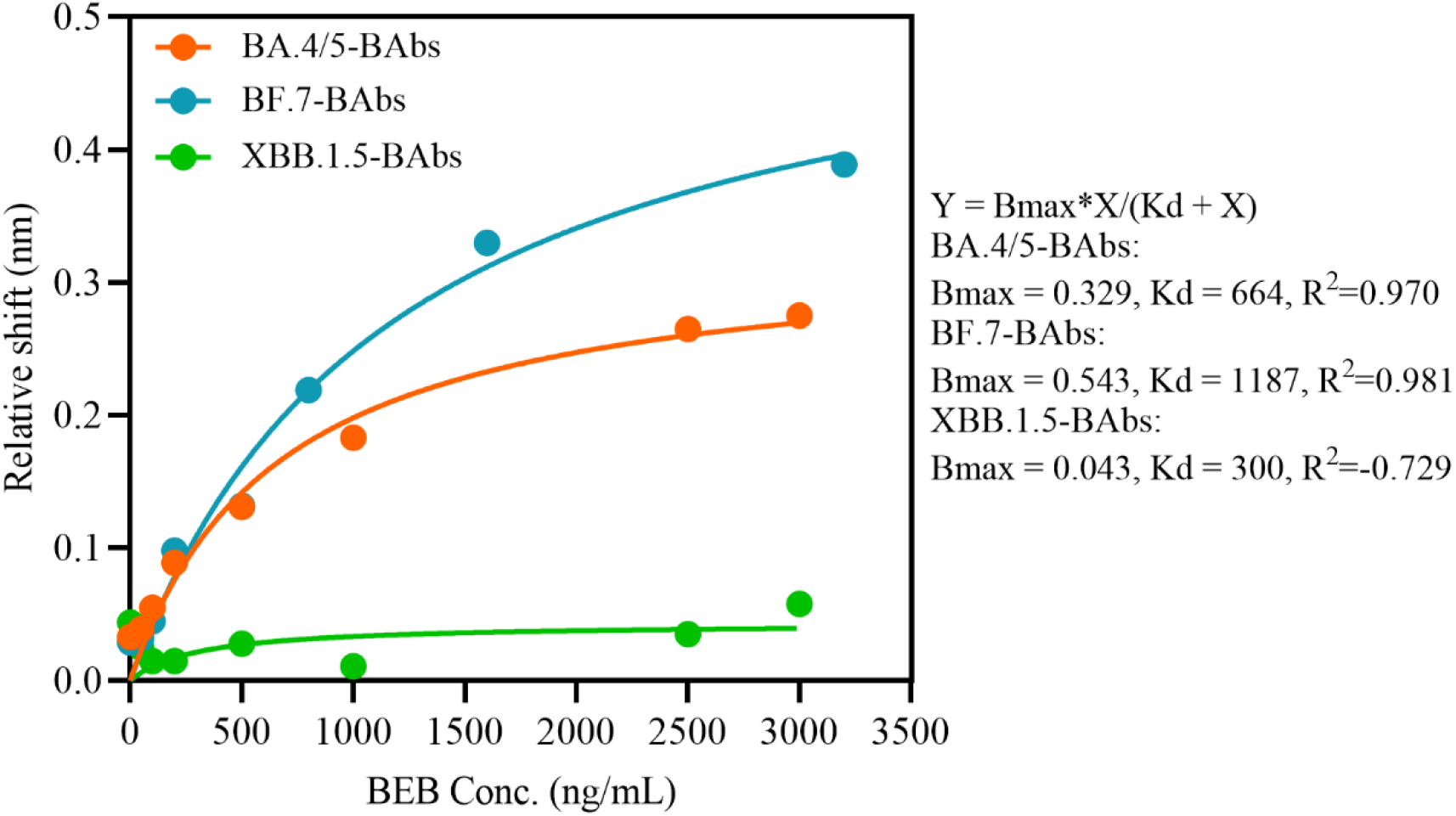
Binding activities of BEB towards BA.4/BA.5, BF.7 and XBB.1.5, respectively, showing no interaction between BEB and the latest XBB.1.5.

### Supplementary Tables

**Table S1.** The FO-BLI biosensor for multiplexed biosensing of NAbs Towards WT and Omicron in Both Sera and DBS Using AMEC as the signal enhancer.

| Main Items | Assay conditions<sup>1</sup> | Anti-WT RBD NAbs | Anti-Omicron RBD NAbs (BA.4/BA.5, BF.7, XBB.1.5) |
| --- | --- | --- | --- |
| **Capture Loading** | Probe | SA sensor | SA sensor |
|  | **Capture protein (conc.)<sup>1</sup>** | **hACE2-B**<br>(5 ug/mL) | **hACE2-B**<br>(5 ug/mL) |
|  | Capture buffer<sup>2</sup> | SD | SD |
|  | Capture shift | 1.0 nm | 1.0 nm |
|  | Washing | 30s in SD | 30s in SD |
| **Sample Processing** | **Sample type** | **Sera, DBS** | **Sera, DBS** |
|  | Sample volume | 5 µL | 5 µL |
|  | Sample dilution | 1/100<br>[DBS: 1/25 pre-diluted] | 1/100<br>[DBS: 1/25 pre-diluted] |
|  | Sample buffer | High-salt SD | High-salt SD |
| **Competitive Binding** | **Protein RBD conjugate** | **WT-HRP** | **Omi.-HRP** |
|  | Protein-HRP conc. | WT-HRP | BA.4/BA.5-HRP: 1/3000<br>BF.7-HRP: 1/4000<br>XBB.1.5-HRP: 1/1000 |
|  | Dilution buffer | High-salt SD | High-salt SD |
|  | Competitive binding | 100 uL sample +<br>100 uL WT-HRP | 100 uL sample +<br>100 uL Omi.-HRP |
|  | Binding time | 5 min | 5 min |
|  | Washing | 30 s in high-salt SD | 30 s in high-salt SD |
| **Amplify Signals** | **Enhancer** | **AMEC** | **AMEC** |
|  | Enhancing time | 1 - 2 min | 1 - 2 min |
| **Assay Properties** | **Calibrators** | **WT-R001** | **BEB (not for XBB.1.5-NAbs)** |
|  | Detection range | 10 - 200 ng/mL | 10 - 2000 ng/mL |
|  | Signal unit | Relative inhibition percentage | Relative inhibition percentage |
|  | LoD for sera | WT-NAbs: 4.9% inhibition | Omi.-NAbs: 0.0% inhibition |
|  | LoD for DBS | WT-NAbs: 0.0% inhibition | Omi.-NAbs: 0.0% inhibition |
|  | **Detection time<sup>3</sup>** | **6.5 - 7.5 min using pre-functionalized fibers** |  |
|  | Fiber regeneration | Not allowed | Not allowed |

<sup>1</sup> All the conjugated protein-HRP obtained an initial concentration of 0.5 mg/mL before use.
<sup>2</sup> The use of SD buffer to dilute hACE2-B reduced the shift drift over time.
<sup>3</sup> NAbs detection can be shortened to 6.5 min by slightly increasing the concentration of protein-HRP conjugate while decreasing the enhancer time to 1 min.
<sup>4</sup> AMEC, 3-Amino-9-ethylcarbazole; BEB, Bebtelovimab; hACE2-B, biotinylated human ACE2; Omi.-HRP, HRP conjugated with omicron RBD protein; SA, Streptavidin; WT, wide type.

**Table S2.** Summary of performance of DBS microsample (A) positive and (B) negative samples in five pre-selected extraction buffers and (C) extraction efficiency of series of DBS spiked samples in the selected buffer. CV, Coefficient of Variation.

(A) Inhibition percentages of one positive sample with spiked concentration at 8 ug/mL (i.e., DBS 8) in five pre-selected buffers compared to their serum counterpart (i.e., NAb 8)
| Sample | Extraction buffer | Inhi.1 | Inhi.2 | Inhi.3 | Inhi.4 | Inhi.5 | Mean | STD | CV |
| --- | --- | --- | --- | --- | --- | --- | --- | --- | --- |
| NAb 8 | 100× sera | 75.5% | 74.0% | 72.0% | 78.8% | N/A | 75.1% | 2.9% | 3.8% |
| DBS 8 | Superblock T20 | 51.9% | 51.8% | 50.2% | 69.5% | 67.9% | 58.2% | 9.6% | 16.4% |
|  | PBS+T80 | 64.8% | 64.9% | 62.6% | 73.1% | 72.6% | 67.6% | 4.9% | 7.2% |
|  | PBS+0.5%BSA | 72.6% | 64.8% | 67.0% | 77.6% | 74.5% | 71.3% | 5.3% | 7.4% |
|  | PBS+0.05%T20 | 64.6% | 73.2% | 68.0% | 79.0% | 77.1% | 72.4% | 6.1% | 8.4% |
|  | PBS+0.05%T20+2%BSA | 70.9% | 71.7% | 77.6% | 80.8% | N/A | 75.2% | 4.8% | 6.3% |

**Table S2 (B)** Inhibition percentages of one negative sample (i.e., DBS 0) in the five pre-selected buffers compared to their serum counterpart (i.e., NAb 0)
| Sample | Extraction Buffer | Inhi.1 | Inhi.2 | Mean | STD | CV |
| --- | --- | --- | --- | --- | --- | --- |
| NAb 0 | 100× sera (as control) | 1.1% | -1.1% | 0.0% | 1.5% | N/A |
| DBS 0 | Superblock T20 | -10.7% | -2.3% | -6.5% | 5.9% | -91.2% |
|  | PBS+T80 | -0.6% | -1.1% | -0.9% | 0.4% | -47.4% |
|  | PBS+0.5%BSA | -0.4% | -1.0% | -0.7% | 0.4% | -59.6% |
|  | PBS+0.05%T20 | 2.8% | 1.5% | 2.1% | 0.9% | 43.3% |
|  | PBS+0.05%T20+2%BSA | 2.9% | 2.5% | 2.7% | 0.2% | 8.9% |

**Table S2 (C)** Extraction efficiency of the DBS microsample using spiked WT-R001 concentrations in the selected extraction buffer of PBS+0.05%T20+2%BSA. Measured concentrations were interpolated from the inhibition curve of the AMEC-based FO-BLI NAb biosensor in sera as shown in Fig. 1B.
| Spiked (ug/mL) | Measured Conc. in DBS extracts (ug/mL, n = 3-6) |  |  |  |  |  |  |  |  | Extraction Efficiency |
| --- | --- | --- | --- | --- | --- | --- | --- | --- | --- | --- |
|  | Test 1 | Test 2 | Test 3 | Test 4 | Test 5 | Test 6 | Mean | STD | CV |  |
| 0 | 0.00 | 0.00 | 0.00 | 0.00 | / | / | 0.00 | 0.00 | N/A | N/A |
| 2.5 | / | 1.98 | 2.15 | 2.21 | / | / | 2.11 | 0.12 | 5.7% | 84.4% |
| 4 | 3.46 | 2.45 | 3.02 | 3.28 | 3.60 | 3.60 | 3.23 | 0.44 | 13.7% | 80.9% |
| 8 | 5.51 | 4.84 | 6.07 | 7.08 | 7.30 | 6.70 | 6.25 | 0.95 | 15.3% | 78.1% |
| 16 | 14.30 | 12.62 | 14.38 | 14.62 | 14.20 | 14.50 | 14.10 | 0.74 | 5.3% | 88.2% |
| Overall extraction efficiency |  |  |  |  |  |  |  |  |  | 80.9% |
| STD |  |  |  |  |  |  |  |  |  | 4.4% |
| CV |  |  |  |  |  |  |  |  |  | 5.4% |

**Table S3.** Seropositivity towards the latest omicron subvariants of all 94 sera samples at all timepoints.

| Sample ID |  | NAbs (in inhibition percentage) |  |  |  |
| --- | --- | --- | --- | --- | --- |
| Time point | Donor Nr. | WT | BA.4/BA.5 | BF.7 | XBB.1.5 |
| Day 7 | 13 | 28.1% | 8.7% | 0.0% | 0.0% |
| Day 14 | 10 | 73.7% | 3.2% | 2.3% | 0.0% |
|  | 11 | 57.7% | 5.0% | 6.5% | 0.0% |
|  | 13 | 36.9% | 9.6% | 0.0% | 0.0% |
| Day 21 | 13 | 42.9% | 8.3% | 0.0% | 0.0% |
| Day 29 | 10 | 60.3% | 6.4% | 0.0% | 0.0% |
| Day 270 | 6 | 4.1% | 3.8% | 9.5% | 0.0% |
|  | 13 | 23.5% | 29.5% | 31.3% | 8.8% |
|  | 14 | 4.3% | 3.0% | 14.9% | 0.0% |

**Table S4.** Summary of the IC50 defined by the PVNT of three sera NAbs towards Omicron subvariants.

| Sample | Subvariants | PVNT (IC50) | FO-BLI (Inhibition) |
| --- | --- | --- | --- |
| S1 | BA.4/BA.5 | 86.57 | 5.0% |
|  | BF.7 | 80.41 | 6.5% |
|  | XBB.1.5 | 0.0% | 0.0% |
| S2 | BA.4/BA.5 | 58.75 | 3.2% |
|  | BF.7 | 37.24 | 2.3% |
|  | XBB.1.5 | 0.0% | 0.0% |
| S3 | BA.4/BA.5 | 409 | 29.5% |
|  | BF.7 | 332.2 | 31.3% |
|  | XBB.1.5 | 134.8 | 8.8% |

**Table S5.** FO-BLI multiplexed biosensing of BAbs towards both WT strain and Omicron in sera.

| Main Items | Assay conditions | Anti-WT RBD<br>BAbs | Anti-Omicron RBD BAbs |  |  |
| --- | --- | --- | --- | --- | --- |
| **Capture Loading** | Probe | SA sensor | SA sensor | SA sensor | SA sensor |
|  | **Capture (conc.)<sup>1</sup>** | **RBD-B**<br>(2 ug/mL) | **BA.4/BA.5-B**<br>(2 ug/mL) | **BF.7-B**<br>(2 ug/mL) | **XBB.1.5-B**<br>(2 ug/mL) |
|  | Capture buffer<sup>2</sup> | SD | SD | SD | SD |
|  | Capture shift | 1.2 nm | 1.2 nm | 1.2 nm | 1.2 nm |
|  | Washing | 30s in SD | 30s in SD | 30s in SD | 30s in SD |
| **Baseline** | **Baseline** | **HCS** | **HCS** | **HCS** | **HCS** |
|  | Matrix dilution | 1/40 | 1/40 | 1/40 | 1/40 |
|  | Baseline time | 2 min | 2 min | 2 min | 2 min |
| **Sample Binding** | **Sample type** | **serum** | **serum** | **serum** | **serum** |
|  | Sample volume | 5 µL | 5 µL | 5 µL | 5 µL |
|  | Sample dilution | 1/40 | 1/40 | 1/40 | 1/40 |
|  | Sample buffer | HS-SD | HS-SD | HS-SD | HS-SD |
|  | Sample time | 5 min | 5 min | 5 min | 5 min |
| **Assay Properties** | **Calibrator<sup>3</sup>** | **BEB** | **BEB** | **BEB** | **N/A** |
|  | Detection range | 50–3000 ng/mL | 50-3000 ng/mL | 50-3000 ng/mL | N/A |
|  | Signal unit | relative binding shift | relative binding shift | relative binding shift | relative binding shift |
|  | Cut-off D | 0.007 nm | 0.014 nm | 0.015 nm | 0.072 nm |
|  | **Detection time** | **7.0 min, using the pre-functionalized probe** |  |  |  |
|  | Fiber regeneration | Allowed |  |  |  |

<sup>1</sup> All the conjugated protein obtained an initial concentration of 1.0 mg/mL before use.
<sup>2</sup> The use of SD buffer to dilute Protein-B reduced the shift drift over time.
<sup>3</sup> Day270 serum of donor 13 may serve as a universal BAbs calibrator for both WT and omicron variants.
<sup>4</sup> BEB, Bebtelovimab; HCS, healthy control serum; HS-SD, high-salt SD buffer.

**Table S6.** Reproducibility of the FO-BLI BAbs biosensor for detecting WT-BAbs in seven individual serum samples from seven individual donors on Day21.

| Donor Nr. | Test 1 (nm) | Test 2 (nm) | Mean (nm) | STD (nm) | CV |
| --- | --- | --- | --- | --- | --- |
| 3 | 0.027 | 0.023 | 0.025 | 0.003 | 11% |
| 9 | 0.092 | 0.079 | 0.085 | 0.009 | 10% |
| 10 | 0.439 | 0.460 | 0.450 | 0.015 | 3% |
| 11 | 0.302 | 0.303 | 0.303 | 0.001 | 0% |
| 13 | 0.137 | 0.137 | 0.137 | 0.000 | 0% |
| 14 | 0.084 | 0.096 | 0.090 | 0.009 | 10% |
| 15 | 0.077 | 0.065 | 0.071 | 0.009 | 12% |

**Table S7.** Accuracy of six regression models to predict the inhibition of Day 29 samples. The three parameters as desired were predicted based on four previous measurements of Days 0–21 and compared to the measured inhibition data.

| Regression Model | Sera NABs (RMSE) | DBS NABs (RMSE) | Sera BAbs (RMSE) |
| --- | --- | --- | --- |
| Linear model – Linear regression | 3.1% | 4.3% | 2.9% |
| Linear model – Lasso | 3.1% | 4.2% | 2.7% |
| Polynomial model – 2 degrees | 4.0% | 4.8% | 3.4% |
| Polynomial model – 3 degrees | 8.1% | 4.6% | 4.9% |
| Support vector machine (SVM) | 11.0% | 7.0% | 9.7% |
| Multilayer perceptron (MLP) | 12.0% | 7.8% | 9.3% |

